# Dysphagia and shortness-of-breath as markers for treatment failure and survival in oropharyngeal cancer after radiation

**DOI:** 10.1101/2022.08.24.22279166

**Authors:** Jarey H. Wang, Lance McCoy, Vivian Salama, Temitayo Ajayi, Cem Dede, Amy Moreno, Abdallah S.R. Mohamed, Katherine A. Hutcheson, Clifton David Fuller, Lisanne V. van Dijk, MD Anderson Symptom Working Group

## Abstract

**Background:** Post-treatment symptoms are a focal point of follow-up visits for head and neck cancer patients. While symptoms such as dysphagia and shortness-of-breath early after treatment may motivate additional work up, their precise association with disease control and survival outcomes is not well established.

**Methods:** This prospective data cohort study of 470 oropharyngeal cancer patients analyzed patient-reported swallowing, choking and shortness-of-breath symptoms at 3-to-6 months following radiotherapy to evaluate their association with overall survival and disease control. Associations between the presence of moderate-to-severe swallowing, choking and mild-to-severe shortness-of-breath and treatment outcomes were analyzed via Cox regression and Kaplan-Meier. The main outcome was overall survival (OS), and the secondary outcomes were local, regional, and distant disease control.

**Results:** The majority of patients (91.3%) were HPV-positive. Median follow-up time was 31.7 months (IQR: 21.9-42.1). Univariable analysis showed significant associations between OS and all three symptoms of swallowing, choking, and shortness-of-breath. A composite variable integrating scores of all three symptoms was significantly associated with OS on multivariable Cox regression (p=0.0018). Additionally, this composite symptom score showed the best predictive value for OS (c-index=0.75). Multivariable analysis also revealed that the composite score was significantly associated with local recurrence/progression (p=0.025). Notably, the same significant associations with OS were seen for HPV-positive only subset analysis (p<0.01 for all symptoms).

**Conclusions:** Quantitative patient-reported measures of dysphagia and shortness-of-breath 3-to-6 months post-treatment are significant predictors of OS and disease recurrence/progression in OPC patients and in HPV-positive OPC only.

## Introduction

Annually, more than 50,000 new cases of head and neck cancer (HNC) are diagnosed in the United States, resulting in an estimated 11,000 deaths^1^. With the decline in overall tobacco usage and the increase of prognostically-favorable human papilloma virus (HPV) related HNC tumors in recent decades, survival and cure rates have increased^2–5^. Better outcomes may also be attributed to improved treatment approaches^6^, which typically involve a combination of surgery, radiation, and chemotherapy.

With improvement survival outcomes, new questions arise regarding optimal strategies for post-treatment evaluation and surveillance. Identification of early indicators (or predictors) of disease recurrence and overall survival is needed to personalize the follow-up of HNC patients more effectively. Use of such predictors could guide surveillance strategies, allowing clinicians to optimize HNC patient care by reducing redundant follow-up procedures (e.g. surveillance imaging) for low risk patients, while intensifying procedures for high risk patients to identify true recurrences early^7,8^.

The majority of studies on HNC treatment outcome prediction have focused on pre-treatment variables, such as tumor information, imaging characteristics and comorbidities^9–14^. In contrast, the predictive value of post-treatment symptoms is less clear, despite the empiric use of patient reported symptoms as indicators of potential recurrence. In other words, even though patients with disease progression or recurrence are often symptomatic^11,15–17^, few studies have provided quantitative survival risk assessment of specific symptoms. For instance, Boysen et al. found that 67% of the recurrences in their cohort were identified through symptom presentation^17^, though the symptom types were not described or their severity quantified. Empirically, locoregional head and neck recurrence can present with trouble swallowing or shortness-of-breath due to tissue infiltration or obstruction. To complicate things, these symptoms are common radiation-induced toxicities, and may indicate a general decline in swallowing muscle functionality (resulting in complications like nutritional deficits and aspiration) rather than indicating tumor recurrence. With the assumption that severe dysphagia from treatment toxicity represents morbid functional issues like aspiration and cachexia, Shune et al. demonstrated that the prevalence of severe dysphagia (i.e. no oral intake status) at any time point was related to overall survival^18^. However, the study did not consider tumor recurrences in their analyses, making it difficult to evaluate the full relationship between dysphagia and survival. It is important to consider that early symptoms of severe dysphagia may be clinically distinct from late symptoms.

Our study investigates whether the patient-reported symptoms – swallowing, choking and shortness-of-breath – at 3-to-6 months after radiotherapy treatment are associated with overall survival (OS) and disease control (local, regional, or distant control) in patients diagnosed with primary oropharyngeal cancer (OPC). We further determine if a combination of symptoms signal unfavorable outcomes. Finally, the stratification value of these symptoms for OS and disease control was investigated for the subpopulation with HPV-positive disease.

## Methods

### Cohort selection and treatment

Patient symptom, tumor, and clinical data were prospectively collected as part an active standardized follow-up registry study that was approved by The University of Texas MD Anderson Cancer Center (MDACC) Institutional Review Board (IRB) under [PA14-0947 data collection, PA11-0809 analysis]. This prospective registry enrolls patients evaluated at MDACC with a suspected or confirmed pathologic diagnosis of carcinoma of the oropharynx, including tonsil, base of tongue (BOT), or squamous cell carcinoma of the head and neck of unknown primary origin. For this study, sequential patients meeting these criteria that received radiotherapy with curative intent between 2015 and 2019 at MDACC were included. Generally, patients were treated with 70 Gy in 33 fractions, most commonly with intensity-modulated radiotherapy (IMRT), intensity-modulated proton therapy (IMPT), or volumetric modulated arc therapy (VMAT). Patients that received previous radiation in the head and neck region (re-irradiated patients) were excluded. Additionally, only patients with known symptoms scores at the 3–6-month follow-up after treatment were included. The patient inclusion diagram is detailed in the **Supplemental Figure 1**.

### MDASI symptom score collection

Symptom scores were systematically collected with the MD Anderson Symptom Inventory-Head and Neck Module (MDASI-HN) questionnaire, as part of the prospective observational cohort registry by the MD Anderson Oropharynx Program Patient Reported Outcomes/Function (PROF) Core^19^. This registry collects clinical, therapy and toxicity information systematically at baseline, weekly during and after radiotherapy (6 weeks, then 3-6, 12, 18 and 24 months). Symptoms in the MDASI-HN are rated on a scale from 0-10, where 0 indicates no subjective symptoms and 10 represents the worst symptom severity imaginable.

### Symptom assessment

The symptoms investigated in this study were: patient-rated shortness-of-breath, choking/coughing (referred to as “choke”), and difficulty swallowing/chewing (referred to as “swallow”) at 3-6 months after completion of radiotherapy. Symptom scores for “choke” and “swallow” were discretized as none-to-mild (MDASI score: 0-5) versus moderate-to-severe (MDASI score: 6-10). This cutoff for dysphagia was previously validated as being an important screening threshold for clinical and quality of life parameters^20^. Alternatively for shortness-of-breath, visual inspection of the OS Kaplan Meyer curves showed that the optimal discriminating threshold was 0-1 (none-to-barely) versus 2-10 (mild-to-severe). It may be that mild shortness-of-breath symptoms are clinically significant for reasons that require additional exploration (as compared to mild dysphagia). Additionally, to assess the value of multiple symptoms together in predicting survival a composite variable of the three symptoms was created based on presence of the symptoms together. Consequently, this composite variable represents the presence of 1, 2, or all 3 of the symptoms based on the discretized thresholds mentioned above.

### Outcome measures

The primary endpoint of this study was overall survival (OS), and the secondary endpoints were local, regional, and distant disease control. The time-to-event was defined from start of definitive RT to time of death (for OS), or to first evidence of progression/recurrence (for disease-free survival) as documented in follow-up visits. Residual tumor shortly after treatment that resolved over time was not considered as a progression or recurrence event.

### Statistical analysis

Univariable and multivariable Cox regression analyses were performed with the following parameters: age, sex, HPV status, T stage, N stage, ECOG performance status, site of tumor (base of tongue, tonsil, unknown primary), three symptoms (shortness-of-breath, choke, and swallow) at both baseline (prior to start of RT) and the 3–6-month follow-up, as well as the composite symptom score at the 3-6-month follow-up. Using the aforementioned thresholds (symptom assessment section), symptoms were discretized in the Cox regression model. Significance was evaluated using the Wald test with threshold set to p<0.05. Multivariable testing was performed on variables that demonstrated p<0.2 on univariable testing to assess for confounding. Performance of variables for predicting survival and disease control was assessed using the concordance index (c-index). For Kaplan-Meier analysis, p-values were calculated using the log-rank test. All statistical analyses were performed using R statistical software (version 4.0.3).

## Results

From oropharyngeal cancer patients with follow-up (**Supplemental Figure 1**), 470 had symptom scores reported at the 3–6-month visit and were included in the analysis. Demographics are shown in **Table 1**. Briefly, mean age at diagnosis was 61.7 ± 9.6 years. Consistent with the OPC population distribution, most patients were male (89.4%), and most had tonsil (43.0%) or base of tongue (48.1%) carcinomas. The vast majority of patient tumors were HPV-positive (91.3%). Only 12 patients died (2.6%), with a median time-to-event of 23.7 months, and 36 patients had progression or recurrence (7.7%). These rates were less frequent than in previously reported HPV-positive cohorts in comparable time-frames^4^. Among the patients who died, 4 deaths were related to early treatment failure (<1 yr), 2 were related to late treatment failure (≥1 yr), and 6 were unrelated to the primary tumor. Median follow-up time for all patients was 31.7 months (interquartile range: 21.9-42.1) after end of radiotherapy.

**Table 1:**
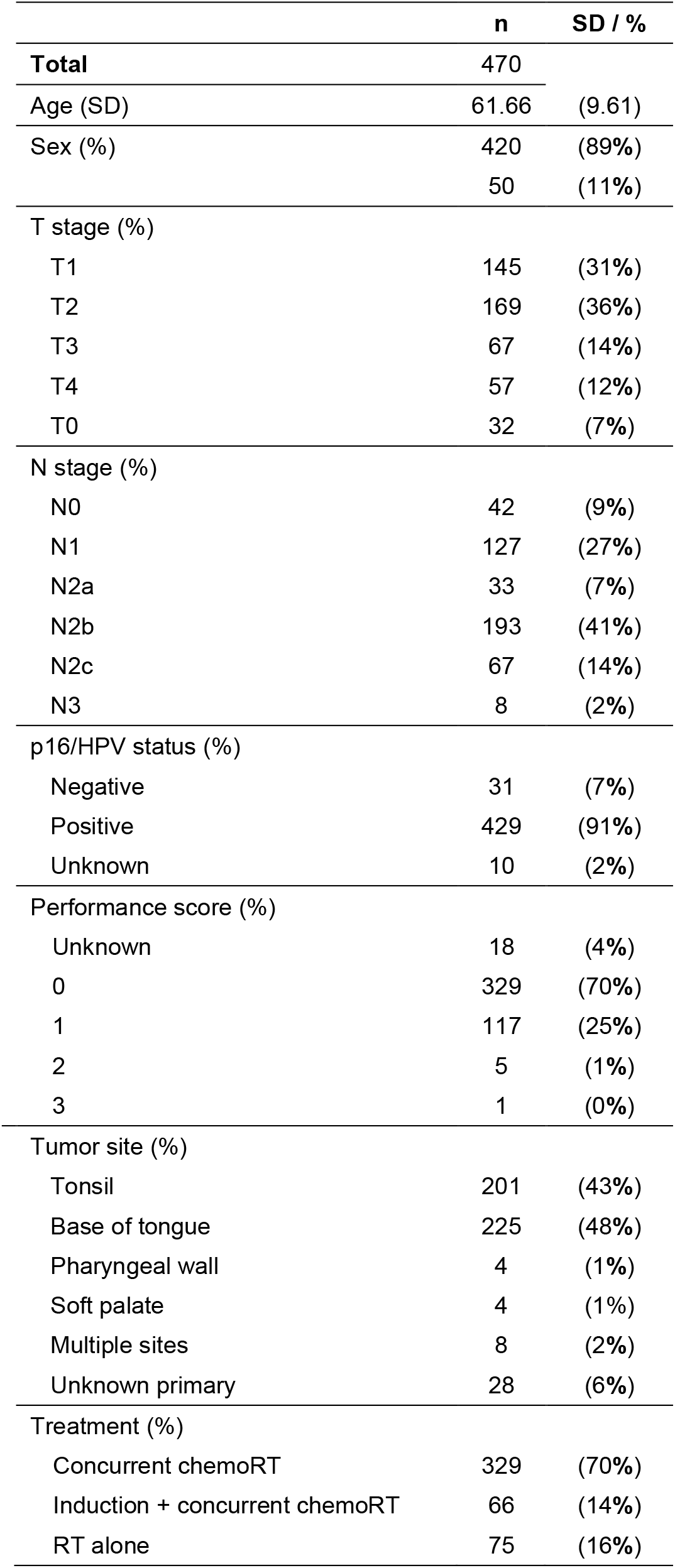
Patient, disease, and treatment characteristics.

|  | n | SD / % |
| --- | --- | --- |
| **Total** | 470 |  |
| Age (SD) | 61.66 | (9.61) |
| Sex (%) | 420 | (89%) |
|  | 50 | (11%) |
| T stage (%) |  |  |
| T1 | 145 | (31%) |
| T2 | 169 | (36%) |
| T3 | 67 | (14%) |
| T4 | 57 | (12%) |
| T0 | 32 | (7%) |
| N stage (%) |  |  |
| N0 | 42 | (9%) |
| N1 | 127 | (27%) |
| N2a | 33 | (7%) |
| N2b | 193 | (41%) |
| N2c | 67 | (14%) |
| N3 | 8 | (2%) |
| p16/HPV status (%) |  |  |
| Negative | 31 | (7%) |
| Positive | 429 | (91%) |
| Unknown | 10 | (2%) |
| Performance score (%) |  |  |
| Unknown | 18 | (4%) |
| 0 | 329 | (70%) |
| 1 | 117 | (25%) |
| 2 | 5 | (1%) |
| 3 | 1 | (0%) |
| Tumor site (%) |  |  |
| Tonsil | 201 | (43%) |
| Base of tongue | 225 | (48%) |
| Pharyngeal wall | 4 | (1%) |
| Soft palate | 4 | (1%) |
| Multiple sites | 8 | (2%) |
| Unknown primary | 28 | (6%) |
| Treatment (%) |  |  |
| Concurrent chemoRT | 329 | (70%) |
| Induction + concurrent chemoRT | 66 | (14%) |
| RT alone | 75 | (16%) |

Of 470 patients, 52 patients (11.1%) presented with moderate-to-severe *swallow* scores (score≥6); 26 patients (5.5%) with moderate-to-severe *choke* scores (score≥6); and 44 patients (9.4%) with mild-to-severe *shortness-of-breath* scores (score≥2). Based on these cutoffs, Kaplan-Meier (KM) curves showed a clear and significant difference in overall survival (OS) for these symptoms (**Figure 1**). When all three symptoms were considered in the composite variable, KM-analysis showed increased mortality for patients for whom all three symptoms were present (p<1e-4, log-rank test) (**Figure 1**).

**Figure 1:**
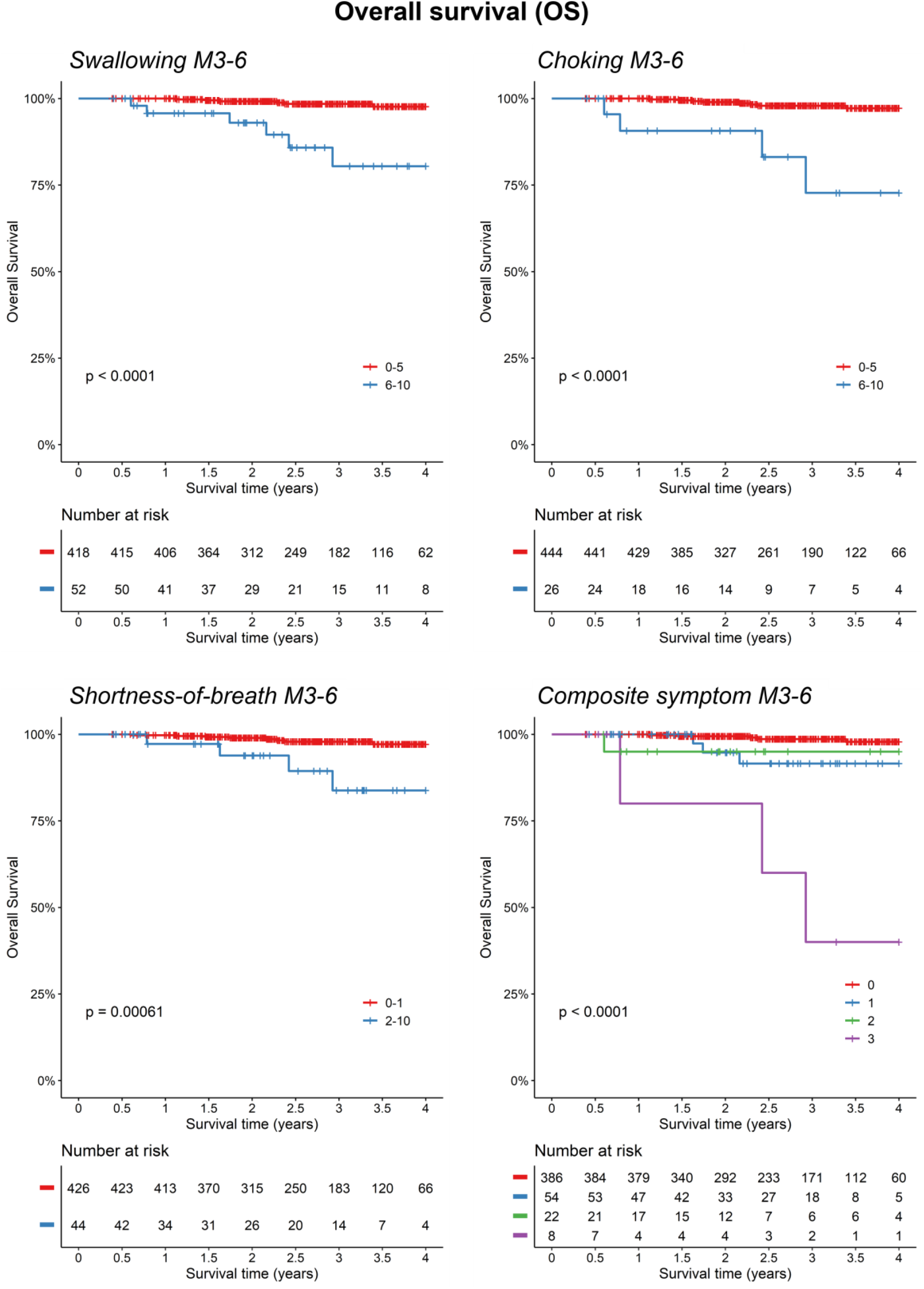
Kaplan-Meier analysis of overall survival (OS) based on patient reported metrics at the 3-to-6 month follow-up. Symptoms of moderate-severe swallowing (MDASI score ≥6), moderate-severe choking (MDASI score ≥6), and mild-severe shortness-of-breath (MDASI score ≥2) at 3-to-6 months post-treatment (M3-6) and a composite symptom score were assessed for association with OS. P-values determined using log-rank test.

Univariable Cox regression analysis for OS (**Table 2**) showed significant associations between OS and 3–6-month MDASI symptom scores for shortness-of-breath (HR=6.26; 95% CI [1.9-21], p=2.9e-3), choke symptoms (HR=11.5 [3.5-38], p=6.7e-5), and swallow symptoms (HR=10.1 [3.3-31], p=6.1e-5). Prediction performance for OS was highest for the swallow score (c-index=0.71 [0.57-0.86]). The composite 3-symptom score outperformed all clinical and individual symptom variables (c-index=0.75 [0.60-0.90]). Two baseline clinical variables were significantly associated with OS in this cohort: tonsil tumor subsite was associated with better OS (HR=0.116 [0.015-0.89]; p=3.9e-2) and age with worse OS (HR=1.06 [1-1.1], p=4.4e-2). Notably, baseline symptom scores were not associated with OS. To assess for confounding, we performed multivariable Cox regression on variables associated with OS with p<0.2 on univariable analysis. On multivariable analysis (**Table 2**), only the composite score remained significantly associated with survival (HR=2.4 [1.38-4.15], p=1.82e-3).

**Table 2:** Univariable and multivariable Cox regression analysis of overall survival (OS) based on baseline variables as well as 3-to-6 month reported symptoms post-treatment.

| OS, univariable | $\beta$ | HR (95% CI) | p.value | c-index | n |
| --- | --- | --- | --- | --- | --- |
| **m6_combined_score\*** | **1.18** | **3.25 (2-5.2)** | **1.20E-06** | **0.747** | **470** |
| **m6_swallow** | **2.32** | **10.1 (3.3-31)** | **6.10E-05** | **0.714** | **470** |
| **m6_choke** | **2.44** | **11.5 (3.5-38)** | **6.70E-05** | **0.651** | **470** |
| **m6_sob** | **1.83** | **6.26 (1.9-21)** | **2.80E-03** | **0.625** | **470** |
| **tumor_tonsil** | **-2.16** | **0.116 (0.015-0.89)** | **3.90E-02** | **0.680** | **470** |
| **age** | **0.0626** | **1.06 (1-1.1)** | **4.40E-02** | **0.719** | **470** |
| n_stage | 1.24 | 3.46 (0.96-12) | 5.80E-02 | 0.643 | 470 |
| tumor_BOT | 1.21 | 3.37 (0.91-12) | 6.90E-02 | 0.642 | 470 |
| p16_hpv_positive | -1.05 | 0.352 (0.076-1.6) | 1.80E-01 | 0.548 | 460 |
| performance | 0.539 | 1.71 (0.64-4.6) | 2.80E-01 | 0.544 | 452 |
| baseline_swallow | 0.824 | 2.28 (0.29-18) | 4.30E-01 | 0.525 | 373 |
| t_stage | 0.119 | 1.13 (0.68-1.9) | 6.40E-01 | 0.553 | 470 |
| sex | 0.326 | 1.39 (0.18-11) | 7.50E-01 | 0.508 | 470 |
| baseline_sob | -0.325 | 0.722 (0.093-5.6) | 7.60E-01 | 0.530 | 378 |
| baseline_choke | -16 | 1.09e-07 (0-Inf) | 1.00E+00 | 0.508 | 375 |

| OS, multivariable | $\beta$ | HR (95% CI) | p.value | c-index | n |
| --- | --- | --- | --- | --- | --- |
| **m6_combined_score\*** | **0.87** | **2.40 (1.38-4.15)** | **1.82E-03** |  | **460** |
| n_numeric | 1.42 | 4.13 (0.74-23.1) | 1.06E-01 |  | 460 |
| tumor_tonsil | -1.51 | 0.22 (0.03-1.84) | 1.63E-01 |  | 460 |
| age | 0.04 | 1.04 (0.96-1.12) | 3.08E-01 |  | 460 |
| p16_hpv_positive | -0.70 | 0.50 (0.11-2.35) | 3.79E-01 |  | 460 |

\*MDASI combined score is an ordinal variable that accounts for all three symptoms: swallow, choke, and shortness of breath.
*Abbreviations:* m6: 3-to-6-month MD Anderson Symptom Inventory-Head and Neck Module; sob: shortness of breath; BOT: base of tongue

Kaplan-Meier analysis also showed significant associations between local control (LC) and swallow (p<1e-4), choke (p<1e-4), and shortness-of-breath scores (p=6.1e-4), as well as for the composite variable (p<1e-4) (**Figure 2A**). Depicted in **Table 3**, univariable Cox regression analyses for LC was significant for swallow (HR=19.5 [3.56-107], p=6.2e-4), choke (HR=11.1 [2.03-60.8], p=5.5e-3), and shortness of breath (HR=6.35 [1.15-35], p=3.4e-2). Prediction performance was highest for the swallow score (c-index=0.82 [0.64-1.00]). The composite 3-symptom score continued to outperform individual variables (c-index=0.84 [0.65-1.00]). T stage was associated with LC as well on univariable Cox analysis (HR=2.36 [1.1-5.03], p=2.7e-2). On multivariable Cox regression (**Supplemental Table 1**), only the composite score remained significantly associated with LC (HR=2.83 [1.14-7.02], p=2.48e-2).

**Table 3:**
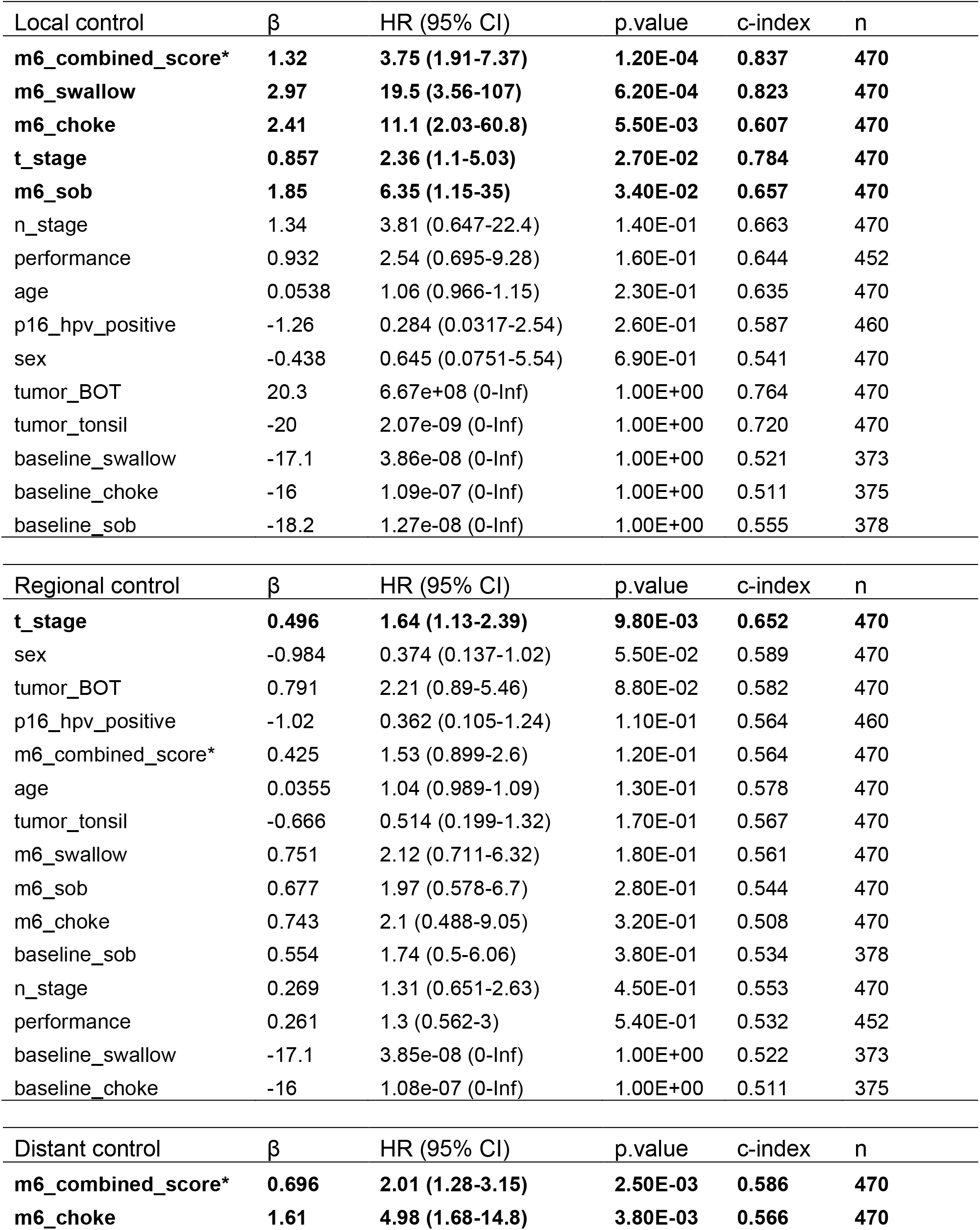
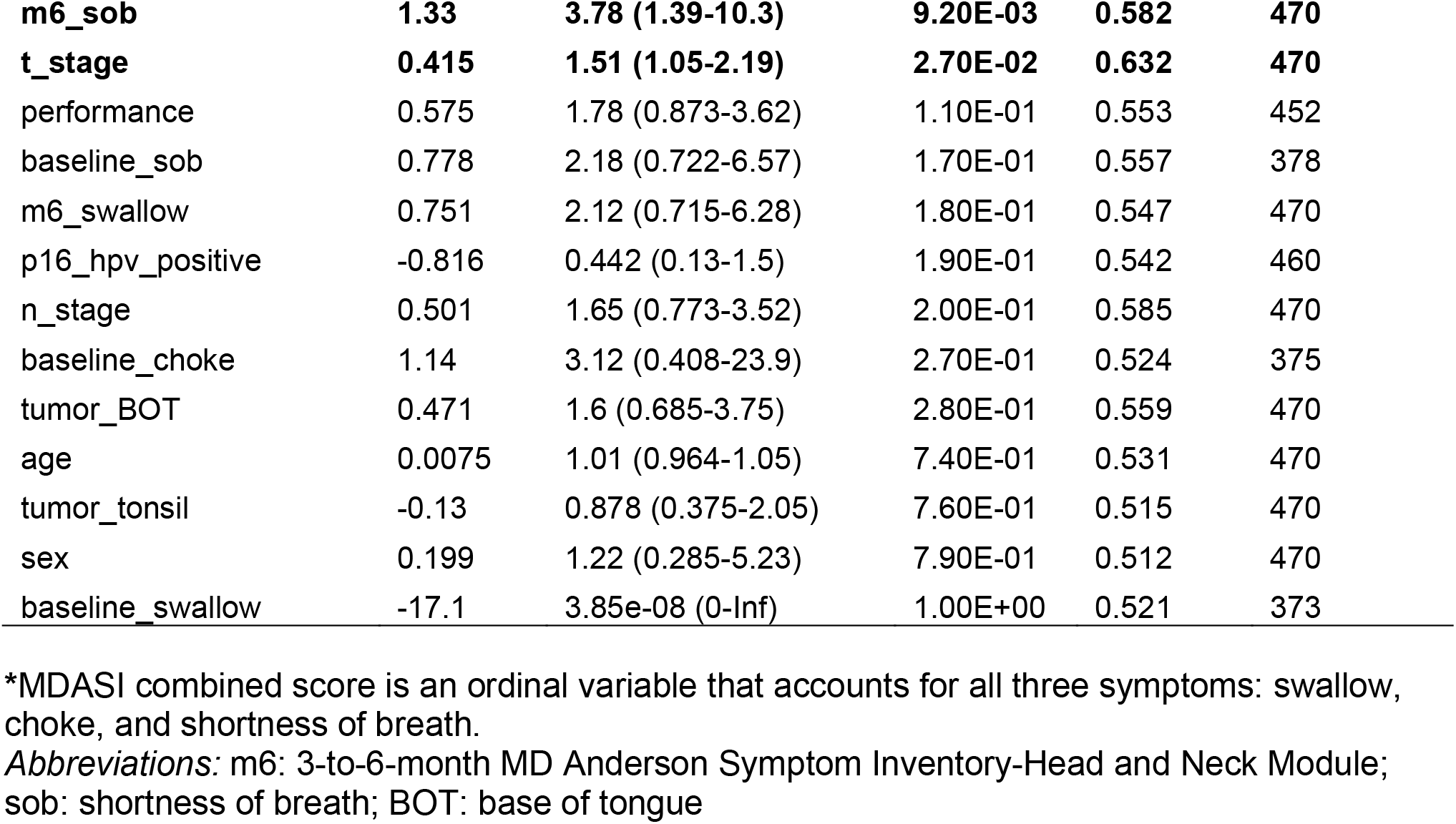
Univariable Cox regression analysis of local, regional, and distant disease-free survival based on baseline variables as well as 3-to-6 month reported symptoms post-treatment.

**Figure 2:**
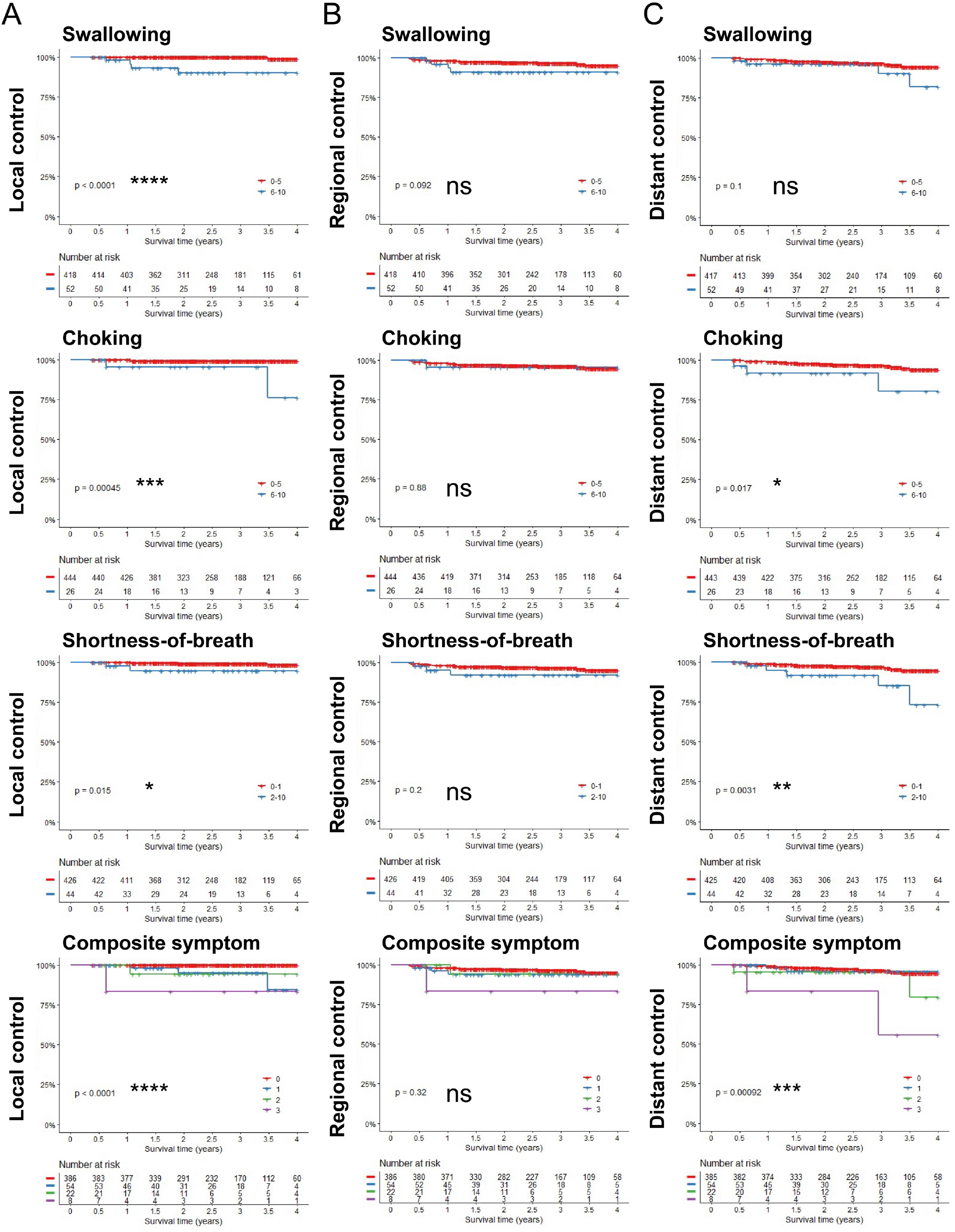
Kaplan-Meier analysis of local, regional, and distant disease-free survival based on patient reported metrics at the 3-to-6 month follow-up. Symptoms of moderate-severe swallowing (MDASI score ≥6), moderate-severe choking (MDASI score ≥6), and mild-severe shortness-of-breath (MDASI score ≥2) assessed at 3-to-6 months post-treatment (M3-6) and a composite symptom score were assessed for association with local, regional, and distant disease control. P-values determined using log-rank test. *p<0.05, **p<0.01, ***p<0.001, ****p<0.0001

**Figure 3:**
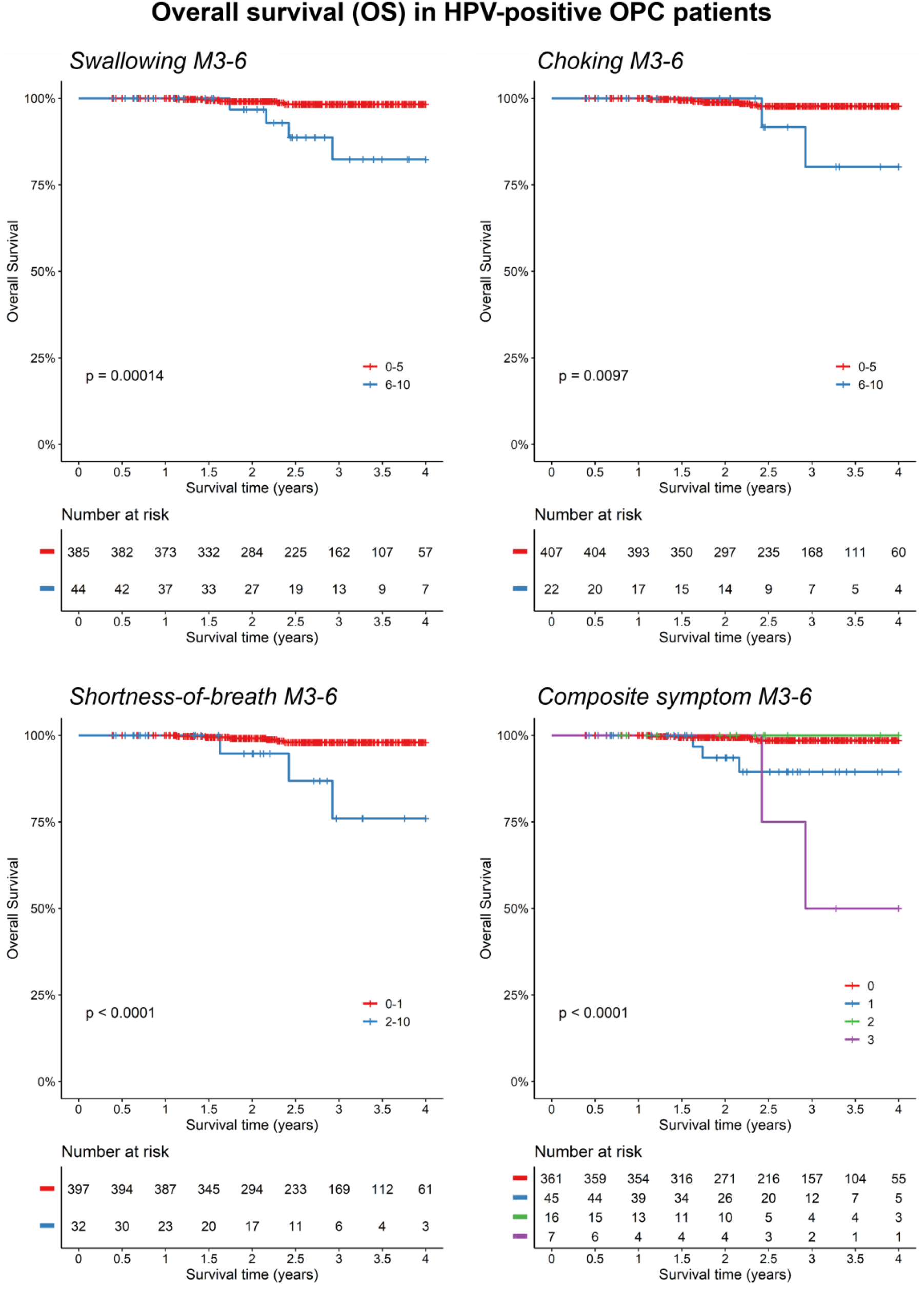
Kaplan-Meier analysis of overall survival (OS) for p16/HPV positive patients based on patient reported metrics at the 3-to-6 month follow-up. Symptoms of moderate-severe swallowing (MDASI score ≥6), moderate-severe choking (MDASI score ≥6), and mild-severe shortness-of-breath (MDASI score ≥2) at 3-to-6 months post-treatment (M3-6) and a composite symptom score were assessed for association with OS. P-values determined using log-rank test.

While symptom scores were related to LC, they were not associated with regional control in KM or Cox regression analyses (**Figure 2B; Table 3**). Disease control for patients with moderate-severe dysphagia scores (swallow, choke) seemed to fall into two categories. For cases of local failure only, the mean latency between end of treatment and failure detection was 30.5 months (SD=12.6). For cases with simultaneously detected local failure and regional and/or distant spread, the mean latency was 9.1 months (SD=3.0).

For distant control (DC), KM-analysis showed significant associations for choke and shortness-of-breath scores (**Figure 2C**). Cox regression corroborated these associations for choke symptoms (HR=4.98 [1.68-14.8], p=3.8e-3) and shortness-of-breath (HR=3.78 [1.39-10.3], p=9.2e-3) (**Table 3**). Having 3 symptoms in the composite variable was associated (p=9.2e-4) with distant failure in KM-analysis (**Figure 2C**, bottom panel), performance on Cox regression (c-index=0.59 [0.47-0.69]) was roughly equivalent to that of shortness-of-breath alone (**Table 3**). For cases of distant failure in patients with mild-severe shortness-of-breath scores, the mean latency to failure from end of treatment was 13.4 months (SD=6.9).

Mortality events that occur after treatment are frequently thought to be associated with disease progression or recurrence. Given the finding that swallow, choke, and shortness-of-breath symptoms were associated with OS and various measures of disease control, we queried whether deaths in the setting of progression or recurrence were associated with more severe symptoms (**Supplemental Table 2**). Progression/recurrence related deaths were significantly associated with moderate-severe swallow (p=2e-3) and choke (p=3e-3) scores, but not associated with shortness-of-breath (p=0.101).

To ensure that these results were not simply reflective of the HPV-negative patients in our cohort, we repeated the analysis with just the 429 HPV-positive patients. Within the HPV-positive group, OS continued to stratify significantly based on symptom severity for swallow (p=1.4e-4), choke (p=9.7e-3), shortness-of-breath (p<1e-4), and the composite symptom variable in KM-analysis (**Supplemental Figure 3**). Analysis of local control (LC) also showed a significant association with swallow scores (p=4.2e-3) and choke scores (p=4.4e-2) **(Supplemental Figure 2A)**. The association between LC and choke symptoms appeared to be driven by events that occurred late (>3 years) after treatment. In contrast to the full patient cohort, shortness-of-breath was not associated with LC in HPV-positive patients. Similar to the full cohort, KM-analysis showed a significant association between distant control (DC) and shortness-of-breath (p=9.7e-3) (**Supplemental Figure 2C**). However, there was no association between DC and choke symptoms.

## Discussion

Along with improved therapeutic approaches, understanding baseline predictors of survival like HPV-status have helped establish HNC care. However, how individual patient factors at post-treatment follow-ups can be used to guide clinical care is less concretely defined and thus a matter of active investigation. This study demonstrates that symptom presentation in the sub-acute setting after radiotherapy can guide current surveillance strategies that are geared toward early detection of treatment failure. We found significant associations between severity of patient-reported swallowing, choking, and shortness-of-breath symptoms at 3-to-6 months after treatment and overall survival (OS), as well as disease control. All three symptoms were significant predictors of OS (**Table 2**), with a composite variable of all three symptoms having the best predictive value (c-index=0.75). Moreover, on multivariable analysis including baseline variables, the composite variable remained the sole significant predictor of OS. Importantly, symptom scores prior to treatment (at baseline) were not significantly associated with outcomes.

These findings are consistent with a finding from Nordgren et al., who reported that subjective swallow scores (based on the EORTC Quality of Life Questionnaire-Head and Neck module) at the 1-year follow-up were significantly different for patients that survived at 5 years compared to those that died in the interim^11^. However, in addition to the different time for symptom reporting, it was unclear whether the observed association was related to recurrence or progression as opposed to treatment side effects^11^.

Our results show that in addition to predicting OS, early symptoms of dysphagia (swallow and choke) are also significantly associated with local disease control, while symptoms of choke and shortness-of-breath are predictors of distant disease control. Specifically, of the 52 patients that experience moderate-to-severe swallowing symptoms (MDASI score≥6), 7.7% had local failure and 11.5% died compared to only 0.5% and 1.4% for both outcomes, respectively, in the patients with no-to-mild swallowing symptoms (**Supplemental Table 2**). While early symptoms of moderate-severe choking were similarly associated with local failure, they were additionally associated with distant metastasis (HR≈5, p=3.8e-3), suggesting that different dysphagia symptoms associate with different outcomes. Shortness-of-breath seemed to be primarily an identifier for distant control (HR≈4, p=9.2e-3).

Previous retrospective studies have reported that most patients with disease recurrence had subjective symptoms at some time prior or during diagnosis of recurrence^11,15–17^. We now establish that already at 3-to-6 months after radiotherapy the presence of moderate-severe dysphagia and mild-severe shortness-of-breath can not only indicate mortality risk, but also risk of treatment failure. Curiously, analyses showed no relation between higher scores of swallow, choke, and shortness-of-breath at the 6-week follow-up and OS (data not presented). This is may be partly due to treatment-induced symptoms having not resolved immediately post-treatment, combined with possible residual tumor. At the 3-to-6-month period acute radiation damage may be resolved, and such severe symptoms then warrant greater concern for recurrence or progression.

Our results also suggest that follow-up symptoms may only portend recurrence in some instances, whereas they may be associated with survival for alternative reasons in other instances. This partly agrees with findings by Shune et al. that mortality can also be caused by the symptoms directly^18^. They argue that a large fraction of HNC patients demonstrating severe dysphagia also exhibit malnutrition, weight loss, and impaired immune function, which often results in cachexia, fatigue, infection risk, or death^21,22^. In contrast to the hypothesis that more severe dysphagia leads to mortality through functional decline, our data suggest that death for patients with more severe dysphagia symptoms (**Supplemental Table 2**) may be frequently cancer-related (swallowing: 66%, 4/6 deaths; choking: 75%, 3/4 deaths). However, symptoms like shortness-of-breath may have more diverse etiologies. Shortness-of-breath may indicate cancer-related pathologies like airway obstruction and pulmonary metastases or may be a more general indicator of decreased performance status. Of the four patients with high shortness-of-breath scores who died, two had pulmonary metastases and two had no evidence of recurrence (**Supplemental Table 2**). In addition, while mild-severe shortness-of-breath was associated with worse overall survival, there was no association between mild-severe symptoms and deaths related to recurrence (p=0.101).

Overall, it is notable that symptom severity – not simply symptom occurrence – is significantly associated with outcomes. Moreover, predictive markers that combine multiple symptoms may be important in ways not previously recognized. This argues for the benefit of quantitative measures of patient reported symptoms during surveillance. Ideally, movement toward standardized, quantitative measures of post-treatment symptoms will help enable the development of more accurate predictive models that will guide surveillance strategies.

The primary limitation of this study is the low number of events, which is likely caused by the high number of patients with HPV-positive OPCs (>90%), which have significantly better prognosis^23,24^. This complicates drawing definite conclusion from our study. More research is needed to verify our results in larger cohort, as well as test the results in a more HPV-negative dominant cohort, which is generally the case in European HNC populations. That being said, the HPV prevalence is a correct reflection of the current OPC population treated at MDACC, which resembles the current incidence of HPV-related OPC in the United States. Importantly, analyses in the HPV-positive OPC cohort showed that our results were not simply reflective of identifying the HPV-negative OPC patients in our study. Notably, HPV status was not significant in univariable analyses, which would likely be different in a larger dataset with more HPV-negative OPCs. An additional limitation is the median follow-up time (31.7 months), which limits analysis of late-onset symptoms. Some studies with extended follow-up have noted the importance of late symptoms as potential indicators of tumor recurrence and toxicities^17,25^.

In conclusion, post-treatment follow-up and surveillance should be tailored to individual needs based on their clinical presentation and symptomology^7,8,16,17^. Our results demonstrate that patient that report individual or combinations of moderate-to-severe swallowing, choking or mild-to-severe shortness-of-breath symptoms as early as 3-to-6 months after treatment have a significantly higher mortality and treatment failure risk. Therefore, more intensified tumor follow-up surveillance may be advised in patients with these symptoms, particularly if all three are present. These results were also applicable in the cohort with only HPV-positive OPC patients.

## Data Availability

All data produced in the present study are available upon reasonable request to the authors

## Author Contributions

Jarey H. Wang: data curation, formal analyses, investigation and methodology, writing (original draft preparation, review, and editing). Lance McCoy: data curation, investigation and methodology. Vivian Salama: data curation, investigation and methodology, writing (review and editing). Temitayo Ajayi: formal analysis, investigation and methodology. Cem Dede: data curation. Amy Moreno: investigation and methodology, resources, writing (review and editing). Abdallah S.R. Mohamed: investigation and methodology, writing (review and editing). Katherine A. Hutcheson: study conceptualization, data curation, investigation and methodology, resources, writing (review and editing). Clifton David Fuller: study conceptualization, supervision, funding acquisition, resources, writing (review and editing). Lisanne V. van Dijk: study conceptualization, data curation, formal analyses, investigation and methodology, funding acquisition, resources, supervision, writing (original draft preparation, review, and editing).

## Data Sharing Statement

The raw datasets that support the findings of this study are not openly available due to identifiable patient information. De-identified portions of the data are available from the corresponding author upon reasonable request.

## Figure Legends

**Supplemental Figure 1:** Inclusion and exclusion criteria for patient selection.

**Supplemental Figure 2:** Kaplan-Meier analysis of local, regional, and distant disease-free survival for p16/HPV positive patients based on patient reported metrics at the 3-to-6 month follow-up. Symptoms of moderate-severe swallowing (MDASI score ≥6), moderate-severe choking (MDASI score ≥6), and mild-severe shortness-of-breath (MDASI score ≥2) at 3-to-6 months post-treatment (M3-6) and a composite symptom score were assessed for association with local, regional, and distant disease control. P-values determined using log-rank test. *p<0.05, **p<0.01, ****p<0.0001

## References

1. Siegel RL, Miller KD, Jemal A. Cancer statistics, 2020. CA Cancer J Clin. 2020;70(1):7–30. doi:10.3322/caac.21590

2. Licitra L, Perrone F, Bossi P, et al. High-risk human papillomavirus affects prognosis in patients with surgically treated oropharyngeal squamous cell carcinoma. J Clin Oncol. 2006;24(36):5630–5636. doi:10.1200/JCO.2005.04.6136

3. Sedaghat AR, Zhang Z, Begum S, et al. Prognostic significance of human papillomavirus in oropharyngeal squamous cell carcinomas. Laryngoscope. 2009;119(8):1542–1549. doi:10.1002/lary.20533

4. Ang KK, Harris J, Wheeler R, et al. Human Papillomavirus and Survival of Patients with Oropharyngeal Cancer. N Engl J Med. 2010;363(1):24–35. doi:10.1056/NEJMoa0912217

5. Mourad M, Jetmore T, Jategaonkar AA, Moubayed S, Moshier E, Urken ML. Epidemiological Trends of Head and Neck Cancer in the United States: A SEER Population Study. J Oral Maxillofac Surg. 2017;75(12):2562–2572. doi:10.1016/j.joms.2017.05.008

6. Lo Nigro C, Denaro N, Merlotti A, Merlano M. Head and neck cancer: improving outcomes with a multidisciplinary approach. Cancer Manag Res. 2017;9:363–371. doi:10.2147/CMAR.S115761

7. Corpman DW, Masroor F, Carpenter DM, Nayak S, Gurushanthaiah D, Wang KH. Posttreatment surveillance PET/CT for HPV-associated oropharyngeal cancer. Head Neck. December 2018:hed.25425. doi:10.1002/hed.25425

8. Ho AS, Tsao GJ, Chen FW, et al. Impact of positron emission tomography/computed tomography surveillance at 12 and 24 months for detecting head and neck cancer recurrence. Cancer. 2013;119(7):1349–1356. doi:10.1002/cncr.27892

9. Baptistella A, Hilleshein K, Beal C, et al. Weight loss as a prognostic factor for recurrence and survival in oropharyngeal squamous cell carcinoma patients. Mol Clin Oncol. October 2018. doi:10.3892/mco.2018.1737

10. Lango MN, Egleston B, Fang C, et al. Baseline health perceptions, dysphagia, and survival in patients with head and neck cancer. Cancer. 2014;120(6):840–847. doi:10.1002/cncr.28482

11. Nordgren M, Hammerlid E, Bjordal K, Ahlner-Elmqvist M, Boysen M, Jannert M. Quality of life in oral carcinoma: A 5-year prospective study. Head Neck. 2008;30(4):461–470. doi:10.1002/hed.20735

12. Quinten C, Maringwa J, Gotay CC, et al. Patient Self-Reports of Symptoms and Clinician Ratings as Predictors of Overall Cancer Survival. JNCI J Natl Cancer Inst. 2011;103(24):1851–1858. doi:10.1093/jnci/djr485

13. Reyes-Gibby CC, Anderson KO, Merriman KW, Todd KH, Shete SS, Hanna EY. Survival Patterns in Squamous Cell Carcinoma of the Head and Neck: Pain as an Independent Prognostic Factor for Survival. J Pain. 2014;15(10):1015–1022. doi:10.1016/j.jpain.2014.07.003

14. Aerts Hjwl, Velazquez ER, Leijenaar RTH, et al. Decoding tumour phenotype by noninvasive imaging using a quantitative radiomics approach. Nat Commun. 2014;5. doi:10.1038/ncomms5006

15. Agrawal A, DeSilva BW, Buckley BM, Schuller DE. Role of the Physician Versus the Patient in the Detection of Recurrent Disease Following Treatment for Head and Neck Cancer. Laryngoscope. 2004;114(2):232–235. doi:10.1097/00005537-200402000-00011

16. Spector ME, Chinn SB, Rosko AJ, et al. Diagnostic modalities for distant metastasis in head and neck squamous cell carcinoma: Are we changing life expectancy? Laryngoscope. 2012;122(7):1507–1511. doi:10.1002/lary.23264

17. Boysen ME, Zätterström UK, Evensen JF. Self-reported symptoms to monitor recurrent head and neck cancer-analysis of 1,678 cases. Anticancer Res. 2016;36(6):2849–2854.

18. Shune SE, Karnell LH, Karnell MP, Van Daele DJ, Funk GF. Association between severity of dysphagia and survival in patients with head and neck cancer. Head Neck. 2012;34(6):776–784. doi:10.1002/hed.21819

19. Rosenthal DI, Mendoza TR, Chambers MS, et al. Measuring head and neck cancer symptom burden: The development and validation of the M. D. Anderson symptom inventory, head and neck module. Head Neck. 2007;29(10):923–931. doi:10.1002/hed.20602

20. Grant S, Kamal M, Mohamed ASR, et al. Single-item discrimination of quality-of-life– altering dysphagia among 714 long-term oropharyngeal cancer survivors: Comparison of patient-reported outcome measures of swallowing. Cancer. 2019;125(10):1654–1664. doi:10.1002/cncr.31957

21. Stephens NA, Skipworth RJE, Fearon KCH. Cachexia, survival and the acute phase response. Curr Opin Support Palliat Care. 2008;2(4):267–274. doi:10.1097/SPC.0b013e3283186be2

22. Gorenc M, Kozjek NR, Strojan P. Malnutrition and cachexia in patients with head and neck cancer treated with (chemo)radiotherapy. Reports Pract Oncol Radiother. 2015;20(4):249–258. doi:10.1016/j.rpor.2015.03.001

23. Vigneswaran N, Williams MD. Epidemiologic Trends in Head and Neck Cancer and Aids in Diagnosis. Oral Maxillofac Surg Clin North Am. 2014;26(2):123–141. doi:10.1016/j.coms.2014.01.001

24. Li H, Torabi SJ, Yarbrough WG, Mehra S, Osborn HA, Judson B. Association of Human Papillomavirus Status at Head and Neck Carcinoma Subsites With Overall Survival. JAMA Otolaryngol Neck Surg. 2018;144(6):519. doi:10.1001/jamaoto.2018.0395

25. Gharzai LA, Li P, Schipper MJ, et al. Characterization of very late dysphagia after chemoradiation for oropharyngeal squamous cell carcinoma. Oral Oncol. 2020;111:104853. doi:10.1016/j.oraloncology.2020.104853

