## Supplementary figures and images for "Dysphagia and shortness-of-breath as markers for treatment failure and survival in oropharyngeal cancer after radiation"

### Supplemental Figure 1

**Supplemental Figure 1: Inclusion and exclusion criteria for patient selection.**

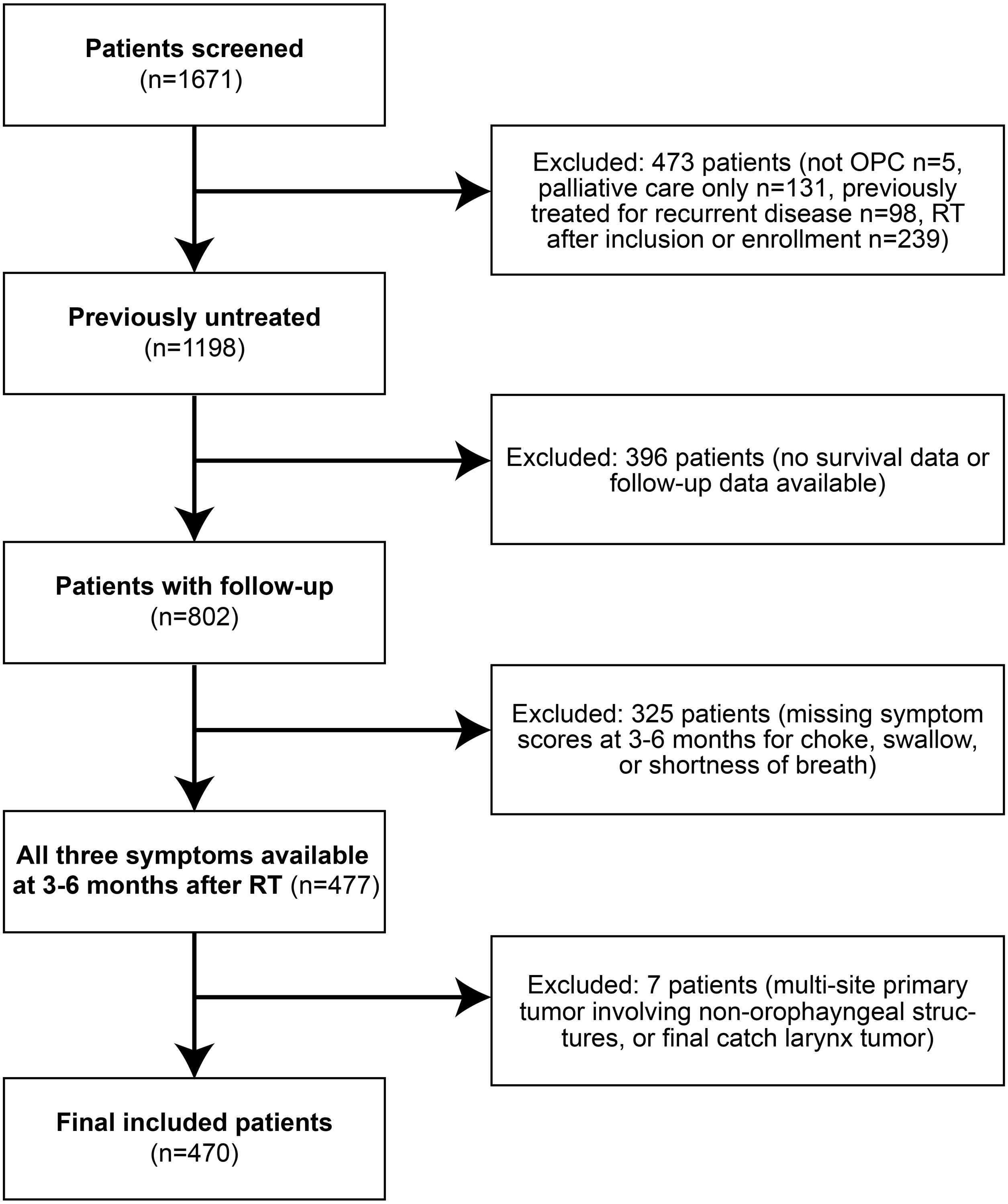
