## Supplemental Figure 2 for "Dysphagia and shortness-of-breath as markers for treatment failure and survival in oropharyngeal cancer after radiation"

**Supplemental Figure 2: Kaplan-Meier analysis of local, regional, and distant disease-free survival for p16/HPV positive patients based on patient reported metrics at the 3-to-6 month follow-up.**

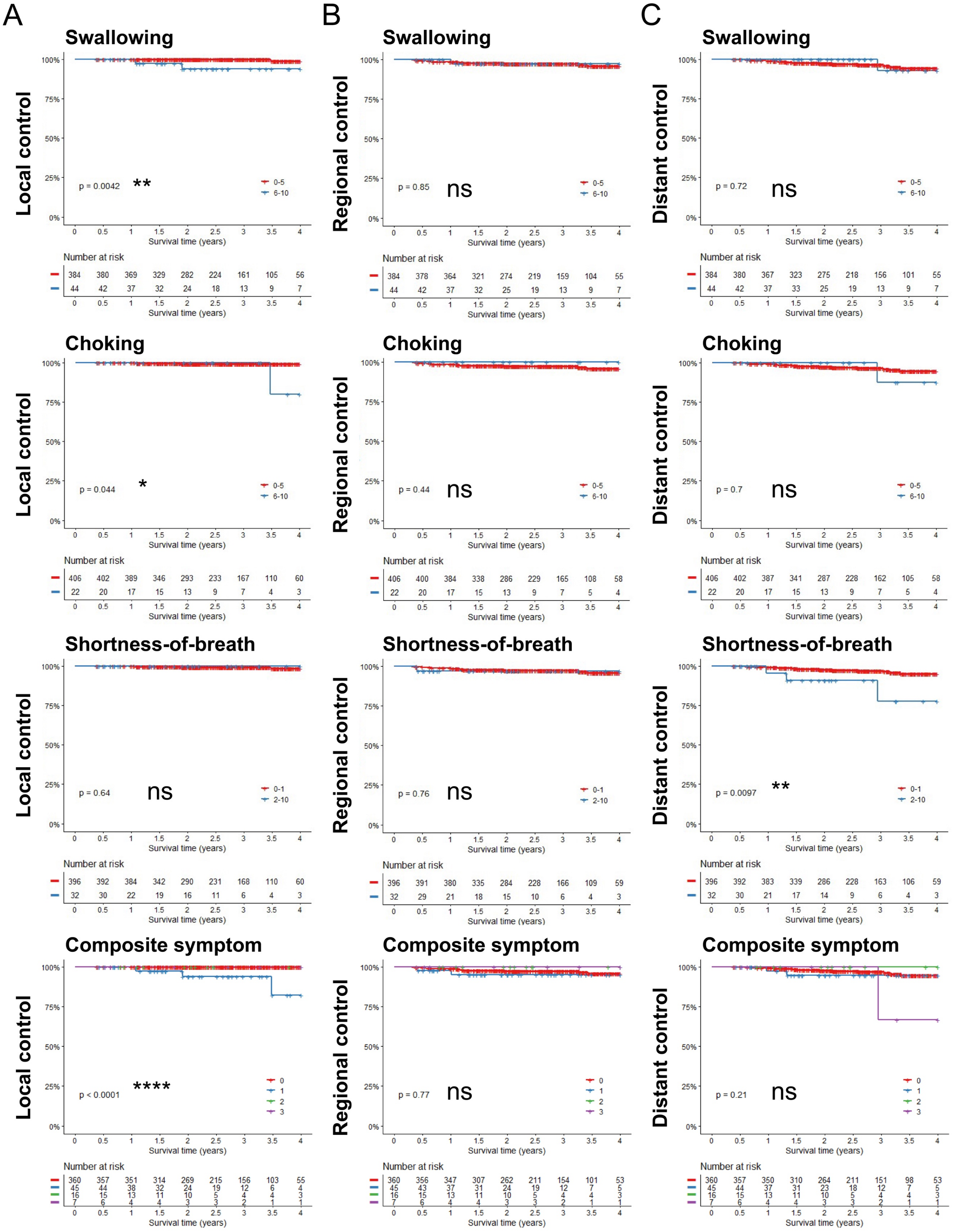
