## Supplemental Tables for "Dysphagia and shortness-of-breath as markers for treatment failure and survival in oropharyngeal cancer after radiation"

**Supplemental Table 1: Multivariable Cox regression analysis of disease-free survival based on baseline variables as well as 3-to-6 month reported symptoms post-treatment.**

| Local control | β | HR (95% CI) | p.value | n |
|---|---|---|---|---|
| **m6_combined_score*** | **1.04** | **2.83 (1.14-7.02)** | **2.48E-02** | **452** |
| t_stage | 0.77 | 2.15 (0.93-4.97) | 7.23E-02 | 452 |
| n_stage | 2.02 | 7.53 (0.38-147) | 1.83E-01 | 452 |
| performance | 0.37 | 1.45 (0.22-9.31) | 6.97E-01 | 452 |

| Distant control | β | HR (95% CI) | p.value | n |
|---|---|---|---|---|
| m6_combined_score* | 0.48 | 1.62 (0.89-2.96) | 0.11 | 359 |
| t_stage | 0.30 | 1.34 (0.86-2.09) | 0.19 | 359 |
| p16_hpv_positive | -0.57 | 0.56 (0.15-2.07) | 0.39 | 359 |
| baseline_sob | 0.44 | 1.55 (0.45-5.35) | 0.49 | 359 |
| performance | 0.02 | 1.02 (0.41-2.54) | 0.97 | 359 |

*****MDASI combined score is an ordinal variable that accounts for all three symptoms: swallow, choke, and shortness of breath.
*Abbreviations:* m6: 3-to-6-month MD Anderson Symptom Inventory-Head and Neck Module; sob: shortness of breath

**Supplemental Table 2: Outcomes for patients with high versus low scores for MDASI for swallow, choke, and shortness of breath.**

| | | Totals (% of total) | Local failure | Regional failure | Distant failure | Deaths | Deaths related to progression or recurrence |
|---|---|---|---|---|---|---|---|
| Swallowing | High | 52 | 4 (7.7%) | 4 (7.7%) | 4 (7.7%) | 6 (11.5%) | 4 (7.7%) |
| | Low | 418 | 2 (0.5%) | 17 (4.1%) | 18 (4.3%) | 6 (1.4%) | 2 (0.5%) |
| | p-value | | 0.002 | 0.273 | 0.288 | 0.001 | **0.002** |
| Choke | High | 26 | 2 (7.7%) | 2 (7.7%) | 4 (15.4%) | 4 (15.4%) | 3 (11.5%) |
| | Low | 444 | 4 (0.9%) | 19 (4.3%) | 18 (4.1%) | 8 (1.8%) | 3 (0.7%) |
| | p-value | | 0.039 | 0.326 | 0.027 | 0.003 | **0.003** |
| Shortness of breath | High | 44 | 2 (4.5%) | 3 (6.8%) | 5 (11.4%) | 4 (9.1%) | 2 (4.5%) |
| | Low | 426 | 4 (0.9%) | 18 (4.2%) | 17 (4.0%) | 8 (1.9%) | 4 (0.9%) |
| | p-value | | 0.101 | 0.434 | 0.045 | 0.019 | 0.101 |
| Composite variable | High | 8 | 1 (12.5%) | 1 (12.5%) | 2 (25%) | 3 (37.5%) | 2 (25%) |

| | | | | | | | |
|---|---|---|---|---|---|---|---|
| | Low | 462 | 5 (1.1%) | 20 (4.3%) | 20 (4.3%) | 9 (1.9%) | 4 (0.9%) |
| | p-value | | 0.098 | 0.308 | 0.049 | 0.001 | **0.004** |

Thresholds for “high” versus “low” MDASI score categories stated: “high” for swallow is a score≥6; for choke, a score≥6; and for shortness of breath, a score≥2. For the composite variable “high” is denoted as having “high” scores for all three symptoms, versus “low” denoting “high” scores for two or fewer symptoms. P-values obtained using Fisher’s exact test.
